# Spatial risk factors for Pillar 1 COVID-19 case counts and mortality in rural eastern England, UK

**DOI:** 10.1101/2020.12.03.20239681

**Authors:** Julii Brainard, Steve Rushton, Tim Winters, Paul R. Hunter

## Abstract

Understanding is still developing about risk factors for COVID-19 infection or mortality. This is especially true with respect to identifying spatial risk factors and therefore identifying which geographic areas have populations who are at greatest risk of acquiring severe disease. This is a secondary analysis of patient records in a confined area of eastern England, covering persons who tested positive for SARS-CoV-2 through end May 2020, including dates of death and residence area. For each residence area (local super output area), we obtained data on air quality, deprivation levels, care home bed capacity, age distribution, rurality, access to employment centres and population density. We considered these covariates as risk factors for excess cases and excess deaths in the 28 days after confirmation of positive covid status relative to the overall case load and death recorded for the study area as a whole. We used the conditional autoregressive Besag-York-Mollie model to investigate the spatial dependency of cases and deaths allowing for a Poisson error structure. Structural equation models were also applied to clarify relationships between predictors and outcomes. Excess case counts or excess deaths were both predicted by the percentage of population age 65 years, care home bed capacity and less rurality: older population and more urban areas saw excess cases. Greater deprivation did not correlate with excess case counts but was significantly linked to higher mortality rates after infection. Neither excess cases nor excess deaths were predicted by population density, travel time to local employment centres or air quality indicators. Only 66% of mortality could be explained by locally high case counts. The results show a clear link between greater deprivation and higher COVID-19 mortality that is separate from wider community prevalence and other spatial risk factors.

## INTRODUCTION

The respiratory illness COVID-19 arises from infection with the SARS-CoV-2 virus. COVID-19 is a still emerging disease that was first identified in early 2020. COVID-19 was declared a *Public Health Emergency of International Concern* on 30 January 2020 (World Health Organization 2020a) and reached pandemic status on 11 March 2020 (World Health Organization 2020b). It is thought that economically damaging and social disruptive measures implemented to control the outbreak and prevent high excess mortality linked to this disease will only cease to be imposed after an effective vaccine and/or a treatment regime is found (Anderson *et al*. 2020).

In high income countries, COVID-19 is thought to have a case fatality rate between 0.2% and 1.5% (Meyerowitz-Katz & Merone 2020, Rajgor *et al*. 2020, Streeck *et al*. 2020). It was clear very early in the outbreak that disease severity was linked strongly to patient age (European Centre for Disease Prevention and Control 2020). COVID-19 is usually a mild illness in children, is not commonly dangerous in adults under 50, and seems to only have mortality rates above 1% in adults above 50 years old (Onder *et al*. 2020). Non-Caucasian ethnicity is also clearly linked to higher hospitalization and mortality rates (Docherty *et al*. 2020). The importance of many other possible risk factors for severe disease outcomes is not as clearly established, however.

This is a secondary analysis of data within a contained region of Eastern England that described persons who tested positive for SARS-CoV-19 using rt-PCR. Residential origin area was available for most patients. This dataset allowed us to explore spatial risk factors that might link to COVID-19 disease incidence and outcomes. We used conditional autoregressive models in the Besag-York-Mollie framework to investigate there was spatial dependence in case detection or mortality following COVID-19 positivity and the extent to which these could be linked to population density, socio-economic deprivation, rurality, levels of air pollution, care home bed capacity, road network connectivity, and age demographics in residential areas.

## METHODS

### DATA

The analysis covers patients who had a confirmed positive swab test by 31 May 2020, and for whom outcomes had been recorded by 22 September 2020. End of May approximately correlated with the end of the first ‘Wave’ of the epidemic in England. A much later extraction date (22 September) than the census date (31 May) was desirable because we knew that it should adequately capture case counts and mortality, rather than have results biased by incomplete data due to delayed reporting.

The dataset described covid+ patients among the population of the English county of Norfolk and a single district (Waveney) in the adjacent county of Suffolk. For historical and geographical reasons, provision of health care in Norfolk and Waveney (N&W) is combined, currently under the commissioning powers held by the N&W clinical commissioning group (NWCCG). Norfolk and Waveney is a predominantly rural and coastal area in Eastern England, UK, that extends roughly 55 by 40 miles. The population is approximately 1 million. Residents of Norfolk are relatively ‘old’ within the UK, with a median age around 46 years which compares to a median age of 40.2 years for all UK residents in mid-2018 (McCurdy 2019). The county is neither especially affluent or deprived but does have areas among the 10% most and least deprived areas in England (Norfolk Insight 2020). Appendix 1 shows percentiles statistics to indicate how representative Norfolk and Waveney are compared to other areas of England for air quality, deprivation, rurality and driving times to nearby employment centres. Deaths among patients with COVID-19 in Norfolk were already known to be strongly linked with advanced age, similar to data from other areas (see data for the single largest acute care provider in N&W in Appendix 2).

The data comprised individual COVID-19 Pillar 1 positive test results for persons in the N&W area who received care from local NHS trusts serving this population. Table 1 lists the NHS trusts who provided Pillar 1 records to NWCCG. Cases detected under the Pillar 1 framework were tested for possible COVID-19 because of medical need for urgent treatment or occupational exposure (Department of Health and Social Care 2020). The dataset did not enable us to separate those tested for medical treatment needs from those with occupational exposure. We believe most of the records relate to persons with medical need, because 57% of the records were for persons age 65 or older (beyond the recent average age of retirement in England; Hofäcker *et al*. 2016), and 75% of the records were for persons age 50+. The data were collected, cleaned and provided to us by NWCCG. The dataset reported which NHS trust identified need for testing, residence area, age, sex, admission date (if applicable), discharge date where applicable, date that COVID-positive swab was taken and date of death when applicable. The data covered patients who tested positive for COVID-19 in the period March 9 to 31 May 2020. 31 May was chosen as the final date because it approximately coincides with the end of the first ‘wave’ in Norfolk, and start of a three month period when local case count was relatively quite low. Fewer than 30 individual patients had multiple positive swab tests, on dates usually within 7 days of each other. We used only the earliest positive test date so that each individual appeared in the dataset only once.

**Table 1.** NHS trusts that provided Pillar 1 test results to NWCCG.

| Acronym | Complete name | Principle types of health care services | #cases start (final) |
| --- | --- | --- | --- |
| ECCH | East Coast Community Health Care | Community hospitals that provide care procedures beyond remit of general practice | 2 (1) |
| JPUH | James Paget University Hospital | Urgent, emergency or consultant-led secondary care | 314 (306) |
| NNUH | Norfolk and Norwich University Hospital | Urgent, emergency or consultant-led secondary care | 890 (870) |
| NSFT | Norfolk and Suffolk Foundation Trust | Mental health and dementia care services | 16 (8) |
| NCHC | Norfolk Community Health and Care | Community hospitals that provide care procedures beyond remit of general practice | 366 (336) |
| Other | Other NHS bodies | Mostly general practice primary care | 392 (240) |
| QEH | Queen Elizabeth Hospital | Urgent, emergency or consultant-led secondary care | 428 (347) |
| WSH | West Suffolk Hospital | Urgent, emergency or consultant-led secondary care | 42 (0) |

Note: #cases start refers to total cases in eligible monitoring period. #cases (final) means total in final analysed dataset. See data selection flowchart in Appendix 4. Some cases were linked to multiple NHS trusts so are counted twice in this table but not in the analysed dataset.

Four acute NHS hospital trusts regularly serve Norfolk and Waveney residents. Data available from NWCCG suggested that historically no more than 3% of NWCCG residents seek acute hospital care from NHS acute care providers not included in our analysis dataset (see information in Appendix 3). All UK residents are entitled to attend any NHS hospital for acute care, but these data help to indicate that most Pillar 1 patients in N&W were likely to be captured in the local NHS trust records available to us. The northern and eastern sides of Norfolk as well as eastern side of Waveney adjoin with the North Sea so NHS care providers in the rest of England are relatively much less accessible (for N&W residents) than the health care providers that are listed in our data.

West Suffolk Hospital (WSH) was unique in our dataset in never providing patient residence information. Omission of these WSH patients could lead to considerable under-ascertainment of COVID-19 cases in areas that had many patients who visited WSH. About 14% of south Norfolk residents preferentially travel to WSH for urgent health care because of shorter driving distances compared to reaching alternative providers (presentation proportions in Appendix 3). Although residential location was missing from the WSH records, the West Suffolk data did list the patient’s primary care provider. Most of the N&W cases linked to WSH (32/42, 76%) were registered with primary care providers in the south Norfolk town of Thetford. Therefore, we excluded the Thetford area (LSOA codes E010264:65-78) from our analysis to better assess any correlations between COVID-19 outcomes and spatial variables for the rest of N&W. The other 10 patients excluded from our analysis because their Pillar 1 test was obtained via contact with WSH were registered with nine different primary health care practices scattered across south and mid-Norfolk.

Home residence for each COVID+ patient was resolved to lower super output area (LSOA). LSOAs are standard census units in England for which socio-economic and other indicators have been calculated. LSOAs are designed to be fairly consistent in population but not geographic size. LSOAs typically each contain about 650 households (OCSI undated) and the 597 eligible LSOAs within N&W each had a median 1567 total residents in mid-2019 (the most recent population estimates available). The information available or possible to derive at LSOA resolution include population density, population counts in specific age groups, rurality/urban characterization, deprivation domains, air quality indicators, transport connectivity. These data allowed us to test whether, for patients who tested positive for COVID-19 and mortality within the subsequent 28 days in each LSOA area could be linked to any of these possible correlative factors, which are described in more detail below.

Counts of persons in specific age ranges in each LSOA was extracted using mid-2019 estimates of persons registered by home address as recorded in general practice (GP) primary care records. These counts were available from the Office of National Statistics (www.ons.gov.uk/peoplepopulationandcommunity/populationandmigration/populationestimates/datasets/lowersuperoutputareamidyearpopulationestimatesnationalstatistics). The supplied data were available in single year bands, from 0 years old to age 90+. We used these data to calculate the percentage of resident population in each N&W LSOA that was age 65 years or older.

Total land area for each LSOA was available from geoportal.statistics.gov.uk/datasets. Combined with the population total in each LSOA, this information enabled us to calculate population density (resident persons/hectare), which we hypothesized might correlate with local physical and social contact rates.

How rural or urban an LSOA was determined by using a classification scheme developed for the Department for the Environment, Food and Rural Affairs, updated to 2014 and available at geoportal.statistics.gov.uk (Bibby & Brindley 2016). The classification schema devised by Bibby & Brindley relied on many decision rules to put areas into one of four categories (from most rural to most urban): hamlet, village, town and fringe or urban depending on the population density, predominant land use in each LSOA and geographical context (such as distinguishing town edges from city edges). The data were available in four ordinal categories (1 = most rural to 4 = most urban) which were handled as a numeric value in the models.

Relative deprivation in each LSOA was indicated by the Index of Multiple Deprivation 2019 (McLennan *et al*. 2019; IMD2019) available from https://www.gov.uk/government/statistics/english-indices-of-deprivation-2019. The IMD2019 is a nationally standardized ranking of relative deprivation encompassing many domains, available for all UK. The values were handled as raw ratio values ranging (within this dataset) from 25 to 32,406. It is more conventional to use IMD2019 values as numeric values indicating relative decile. However, this was undesirable in this dataset because data resolution would have been lost; most LSOAs in this study area are in IMD2019 deciles 3-7.

Air quality indicators were available for each LSOA as subdomain information within the IMD2019. The air quality measures were reported as concentration scores relative to (national standards for hazardous) reference thresholds (McLennan *et al*. 2019). Air quality was reported in five possible domains: NO_2_, SO_2_, benzene, particulates and sum of the preceding four. Appendix 1 shows percentile distributions of air quality indicators (particulates and total scores) for N&W relative to the national distribution for all England. N&W LSOAs are fairly representative of national population exposures for SO_2_, benzene and particulates. N&W has better total air quality and lower NO_2_. These values were highly inter-correlated with each other which made it inappropriate to put them all in one model. While SO_2_ concentrations varied little within N&W (Appendix 1), preliminary research has linked higher concentrations of fine particulate matter with higher COVID-19 mortality in the United States (Wu *et al*. 2020). Therefore, we tried in the modelling only two of the air quality measures as potential correlates: particulate levels and total air quality score.

As of mid May 2020, Norfolk was one of the English counties with fewest COVID-19 cases and deaths (Drury 2020, Place 2020). This low incidence was posited to relate to poor transport and infrastructure links to reach Norfolk from the rest of the UK. We hypothesized that poor transport links might also account for some variation in case counts between N&W LSOAs. Therefore, we obtained calculations of transport connectivity in 2017 available at LSOA level for the Department of Transport and supplied at www.gov.uk/government/statistical-data-sets/journey-time-statistics-data-tables-jts#journey-times-connectivity-jts09. The JTS09 dataset report on many modes of transport but it was inappropriate to put more than one of the measures into a single model due to high multi-collinearity. From this dataset we tested only the variable “Travel time in minutes by car to nearest employment centre with 500-4999 jobs” as a connectivity indicator because N&W is a predominantly rural area with a single main city (Norwich: mid-2019 population estimate = 150,000) and many smaller market towns (typical population 2000-13,000). Private motor vehicles are known to be the main mode of transport in N&W, tending to account for more than 60% of journeys (Norfolk County Council 2016, Great Yarmouth Borough Council 2019).

During the first wave of COVID-19 in England, it was widely acknowledged that a high proportion of deaths were among persons living in residential care homes (Burki 2020). We obtained information about the total number of social care homes in each LSOA and their bed capacity from the Care Quality Commission (https://www.cqc.org.uk/about-us/transparency/using-cqc-data). These counts were each also considered as potential predictors of COVID-19 cases or deaths.

Notably, our dataset does not contain information about ethnic profile of the population within each LSOA. Although such data are available within LSOAs, N&W is an area that is quite low in ethnic diversity, and this is even truer among persons who are most at risk of hospitalization or death from COVID-19 (age 65+). 96.5% of all-age Norfolk residents self-identified as ‘White’ in the 2011 Census (Norfolk Insight 2020). There was negligible utility in trying to test whether percentage BAME persons in residence areas might be predictive of COVID+ outcomes within this population. We view this omission as a strength as much as it is a limitation because relationships with correlates can be interpreted without uncertainty about whether minority ethnic demographics interacted differently with each other correlate.

### ANALYSIS

The spatial nature of the data meant that models had to account for possible spatial autocorrelation. Hence, we used the Besag-York-Mollie model (BYM) which is a conditional autoregressive model (Besag *et al*. 1991) to model the excess number of cases and deaths in LSOAs relative to those expected given the population size in each LSOA and total number of cases, deaths and resident population of the entire study area. The BYM model is autoregressive in that it assesses the contribution of cases and deaths in neighbouring LSOAs, to those recorded in individual LSOAs, based on geographical contiguity. The socio-demographic features for each LSOA were fixed effects in our models. We used these attributes as predictors in ecological regression to investigate their contribution to excess cases and excess deaths across the LSOAs, in comparison to the incidence of cases or deaths in the full study area. We analysed excess cases and deaths with an initial model using all predictors and then refined the model to one in which all predictors had a significant impact on explaining risk. Variance inflation factors were calculated to confirm that coefficient confidence intervals were not excessively biased by multi-collinearity.

To summarise, we tested in each LSOA whether for cases diagnosed by end May 2020, and compared to the full study area, the selected spatial variables correlated with

i. Excess risk of cases in LSOAs, ie Expected cases = pop of LSOA * (study area total cases/study area total population)
ii. Expected deaths = Same area cases * (study area total deaths /study area total cases)

Preferred candidate models had the lowest Deviance Information Criterion for the BYM intercept (Berg *et al*. 2004). Inclusion of variables in the final models was decided using the confidence intervals for parameter estimates (the regression coefficients). Variables for which the 95% confidence interval (95% CI) did not include zero were considered significant. A coefficient 95% CI above zero indicates significant prediction of excess deaths; below zero 95% CI would mean reduction in deaths compared to study area as a whole. Models were fit with a Poisson error structure. Models with zero-inflated structure for count data (ZI Poisson models) were applied because they allow for structural effects. Ten per cent of LSOAs had no cases, more than 60% had no deaths. The total variation explained by the spatial distribution of LSOAs was noted to check if there was spatial clustering. Analyses were undertaken in R using the INLA package.

Some covariates were not independent of each other. To investigate possible relationships between covariates and the case and mortality responses we used Structural Equation Modelling (*Rushton et al*. 2010). It was likely that the socio-demography of the population at risk depended on where people lived, with urban LSOAs having different population structure to those in rural areas. We created a conceptual model of the interactions between variables as a path diagram and then challenged the model with the observed data for the 597 LSOAs. We used a principle of parsimony, removing non-significant pathways from the model, to identify the suite of pathways that best represented relationships in the data, quantifying indirect pathways that impacted on both the number of cases and the number of deaths in LSOAs. Models were fit using the piecewise SEM package in R.

## RESULTS

1977 unique individual patient records were eligible, that reported on confirmed swab tests by 31 May 2020 and who received their test result from contact with at least one of the eligible N&W health care trusts. Appendix 4 shows the procedure for subsetting eligible patients from the full dataset supplied.

Figure 1 shows % of resident population age 65+ for each LSOA mapped in quintiles with main town and city names, while Figure 2 shows deprivation mapped in quintiles groups. In this study area, urban centres are relatively less prosperous and have a younger population than surrounding suburbs.

**Figure 1.**
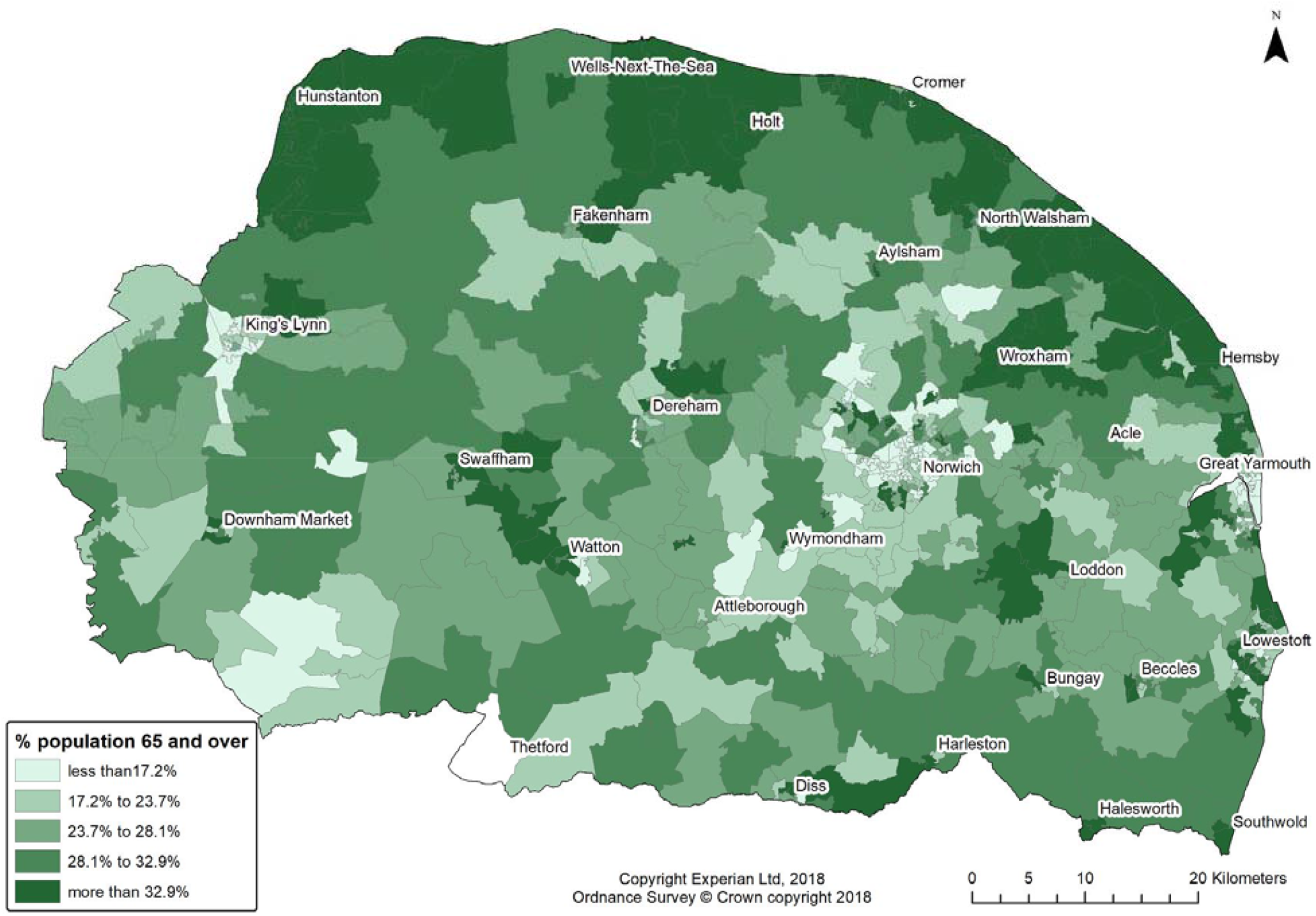
Proportion of population in each LSOA that was age 65 years and over

**Figure 2.**
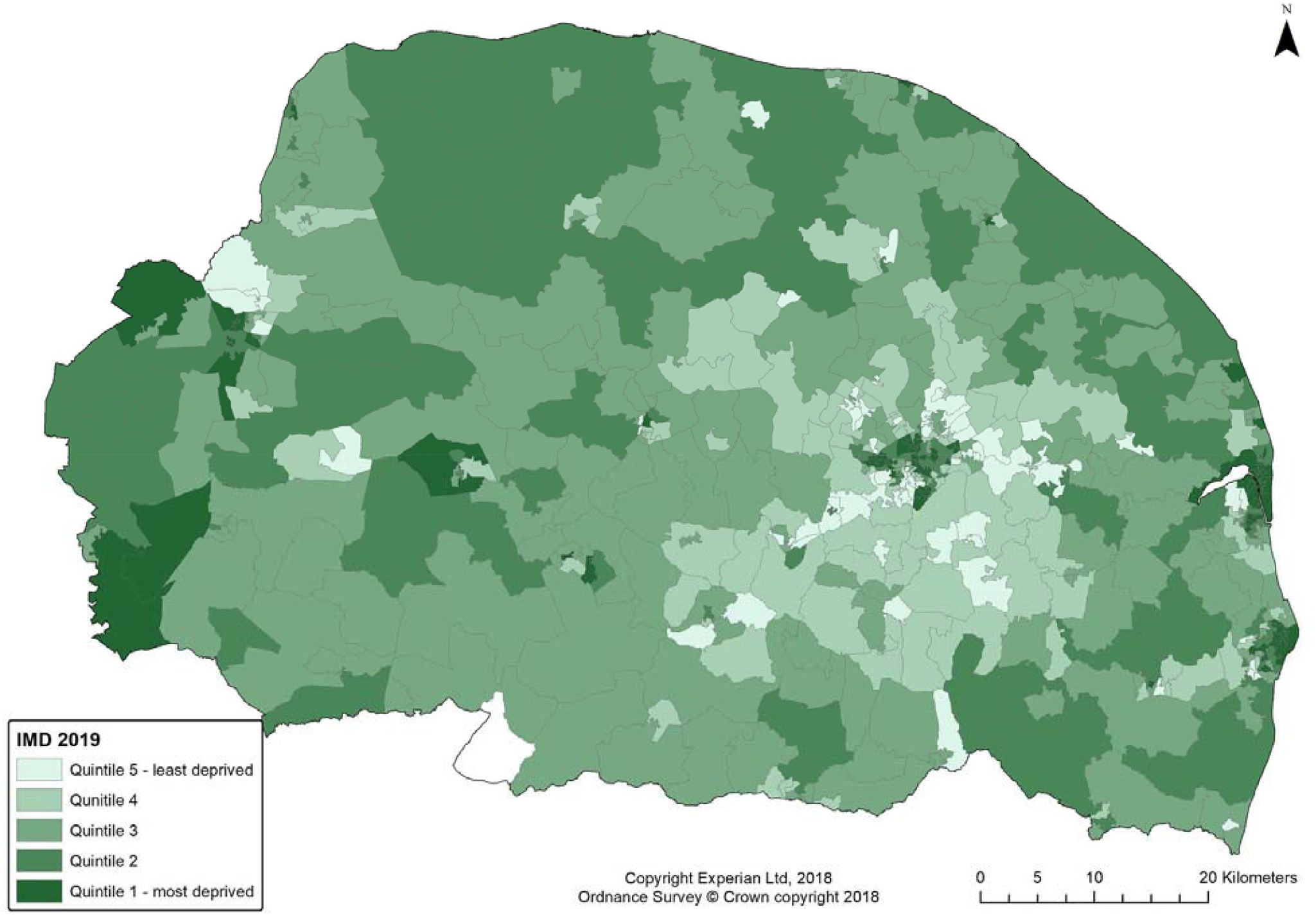
Index of Multiple Deprivation 2019 quintiles in study area

Results were

### i) Risk of excess cases

Three variables significantly predicted excess cases of COVID-19 in LSOAs. These were proportion of the LSOA resident population that was greater than 65 years of age (coefficient estimate=1.193, 95% CI: 1.403-1.978) greater urbanness (estimate 0.133, 95% CI: 0.053-0.214) and care home capacity (estimate 0.016, 95%CI 0.013-0.019). These indicate that aged populations in more urban areas or areas with more care homes had an excess of cases relative to LSOAs with younger populations in more rural settings. Analysis of the proportion of variation explained by the final model was low at 0.01 (on a scale of 0-1, where 1 is maximum possible variance explained by spatial distribution).

### ii) Risk of excess deaths

Three variables were predictors of excess risk of dying after allowing for total cases in the same LSOA; the proportion of the population over 65 (estimate 3.632, 95%CI: 2.281-4.994), the care home bed capacity (estimate 0.028, 95%CI: 0.023-0.033) and lower deprivation for the LSOA (estimate −0.331, 95%CI:−0.506 to −0.160). This indicates that risk of dying was dependent on the proportion of the population over 65 and the extent of deprivation, with more deprived LSOAs having higher excess mortality, as well as local concentration of care home provision. The proportional contribution to the total variation explained by the spatial distribution of LSOAs was 0.02.

Variance inflation factors for both models were all below 1.2, indicating collinearity did not bias model coefficients (Appendix 6).

#### Structural Equation Models

Figure 3 represents covariance between the candidate predictors and each other or the dependent variables used in our models, as obtained from SEMs. Table 2 shows the full model results. The numbers are the standard deviation change at the end of the arrow arising from one standard deviation change in the (origin) predictor. For example, a one standard deviation change in deprivation (IMD) leads to a 0.21 change in standard deviation change in the population % over age 65 variable. The greater the number, the stronger the covariance. These data indicate that increase in case counts explained 66% of the increase in mortality within LSOAs. There was clear indication that the covariates used in the conditional autoregressive models analyses were not independent of each other. The percentage of the population age 65 years and older was especially likely to correlate with other spatially measured factors.

**Table 2.** Selection of structural equation model coefficient results

| Response | Predictor | Estimate | Std.Error | DF | Critical Value | P.Value | Std.Estimate |
| --- | --- | --- | --- | --- | --- | --- | --- |
| Cases | % age 65+ | 4.6864 | 1.6597 | 592 | 2.8236 | 0.0049 | 0.1111 ** |
| Cases | Rurality | 0.3804 | 0.1579 | 592 | 2.4085 | 0.0163 | 0.0911 * |
| Cases | Σ population | 0.0021 | 0.0003 | 592 | 6.7509 | 0.0000 | 0.2519 *** |
| Cases | CH capacity | 0.0777 | 0.0073 | 592 | 10.6997 | 0.0000 | 0.3881 *** |
| Died | Cases | 0.2264 | 0.0101 | 592 | 22.4243 | 0.0000 | 0.6622 *** |
| Died | % age 65+ | 1.8601 | 0.4001 | 592 | 4.6493 | 0.0000 | 0.1290 *** |
| Died | IMD | -0.1830 | 0.0463 | 592 | -3.9513 | 0.0001 | 0.1090 *** |
| Died | CH capacity | 0.0099 | 0.0020 | 592 | 4.8828 | 0.0000 | 0.1447 *** |
| % age 65+ | Rurality | -0.0287 | 0.0038 | 593 | -7.6322 | 0.0000 | 0.2902 *** |
| % age 65+ | CH capacity | 0.0005 | 0.0002 | 593 | 2.5906 | 0.0098 | 0.0979 ** |
| % age 65+ | IMD | 0.0248 | 0.0044 | 593 | 5.6013 | 0.0000 | 0.2130 *** |

Notes: CH capacity = care home bed capacity. Died = covid+ patients who died within 28 days of +swab. DF = degrees of freedom. IMD = Index of Multiple Deprivation 2019. Std = standard. The rurality and IMD (deprivation) scales are such that low rank are more rural/more deprived. Significance thresholds ( $p <$ ): \* 0.05, \*\* 0.01, \*\*\* 0.001.

**Figure 3.**
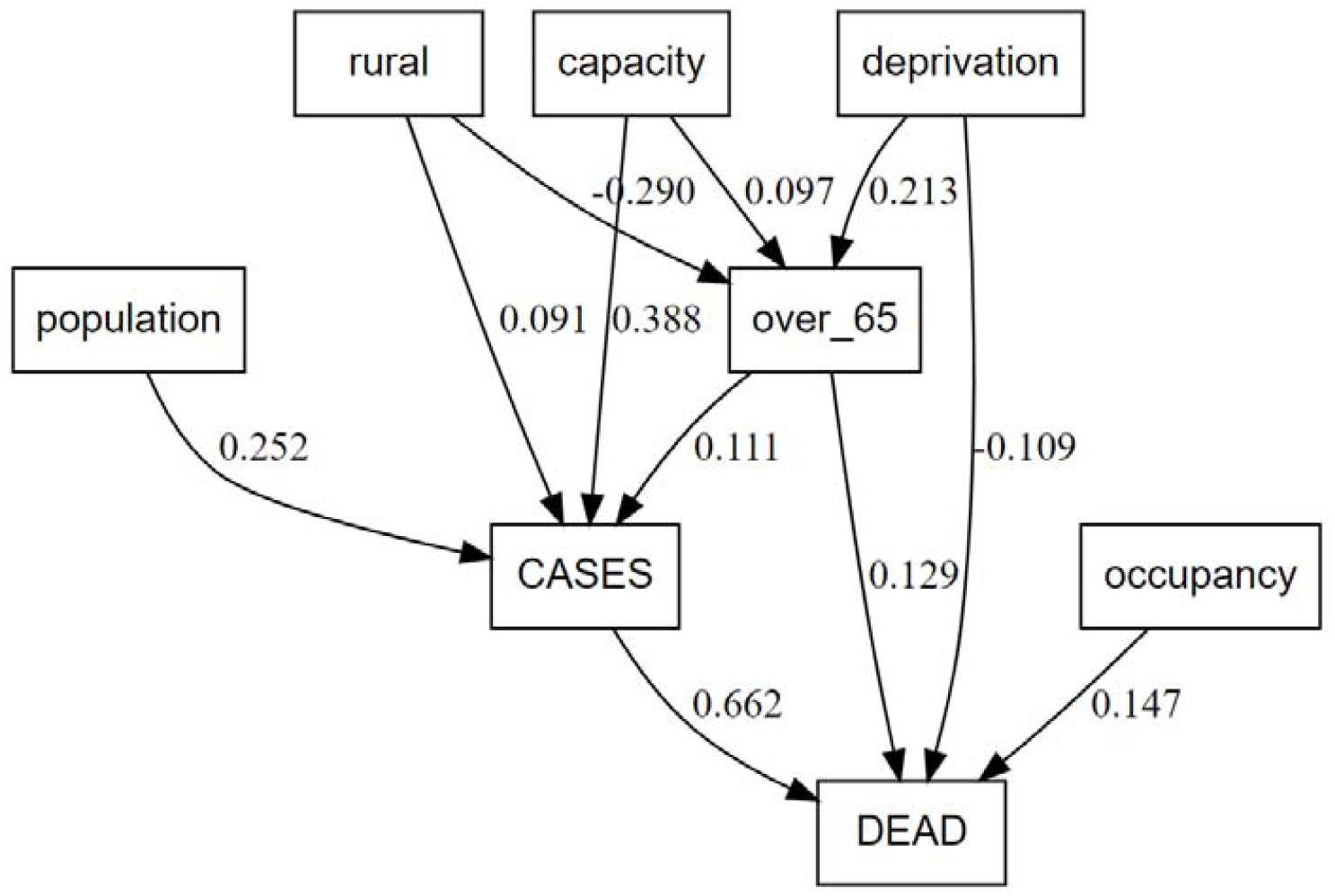
Summary statistics for structural equation (pathway) models

The SEM analyses show that the number of cases in an LSOA was significantly related to higher proportion of population over 65 and lack of rurality of the LSOA. However, the standardized coefficients were below 0.1 indicating that these parameters explained less than 10% of the variation in the number of cases. The number of cases was also strongly related to bed capacity in care homes in LSOAs, effectively representing the population at greatest risk. The number of deaths was strongly related to the total number of cases by end May, as well as greater proportion of the population over 65, higher deprivation and greater care home bed capacity.

## DISCUSSION

Higher case counts in areas with a higher proportion of residents age 65+ probably reflects higher probability of severe disease (given that severity of disease is linked to advanced age). People tend to associate with persons close to their own age and a higher density of older adults may mean more social contacts between older adults with each other (Jackson & López-Pintado 2013). That worse outcomes (higher mortality) were linked to higher deprivation was not surprising. A link between worse COVID-19 outcomes and lower socio-economic status has been reported in other COVID-19 research (Lewis *et al*. 2020, Raisi-Estabragh *et al*. 2020). That more socio-economically deprived areas tend to suffer worse in pandemics may be a common feature of novel respiratory disease outbreaks, and was also a feature of the 2009 influenza pandemic in England and Wales (Rutter *et al*. 2012).

Finding increased mortality with less rurality probably reflects greater mixing and expanded social contact networks for those who live within urban areas, and closer links to outside higher incidence areas. Lower mortality with increased rurality was also observed during the 1918-19 influenza pandemic in England Wales (Chowell *et al*. 2008). However, higher rurality is an unreliable possible protection; arrival of pandemic conditions may only be delayed rather than prevented by rurality. For instance, the second wave of pandemic influenza affected rural areas much worse than the first outbreak wave had done in Wisconsin in 2009 (Truelove *et al*. 2011).

Negative findings (lack of correlation) in our models are useful results. Relative transport accessibility as indicated by the journey times measure did not emerge as significant predictor of cases or deaths in our models. Nor were air quality indicators predictors of case count or mortality. Population density had no separate effect on case count or deaths.

We note that the spatial variation in cases and deaths was negligible. There was little spatial dependency in the number of cases in LSOAs: neighbouring LSOAs did not influence numbers of cases in adjacent LSOAs. Pillar 1 cases and subsequent deaths were not highly clustered at LSOA level geography. Cases and deaths seemed to occur independently of observable spatial contiguity.

### Limitations

The findings relate very much to the types of cases that are found under the Pillar 1 testing framework. These are positive swabs found by testing health professionals (often found through surveillance rather than symptomatic presentation) and patients with urgent medical need. A more conventional sampling framework could include all symptomatic cases (including those without urgent medical needs). Some such cases were found concurrently in May 2020 under Pillar 2 testing protocols. Possible demographic differences between Pillar 1 and Pillar 2 cases in the county of Norfolk alone are described in the data shown as Appendix 5. Pillar 2 and Pillar 1 patients were not very different from each other in simple demographic traits (age distribution and sex).

We have only incomplete information about the occupations of COVID+ patients in our dataset; occupational risk may well have been more important than anything to do with residential origin for individual case status. However, most cases were above age 60 while most deceased were older than the statutory pension age (67 years currently). Occupational exposure is especially unlikely to be relevant to the mortality outcome.

We have tried to be transparent about the covariate specifications. We do not believe that different thresholds (such as considering population age 70 or older) would change the broad conclusions. Lack of variation in ethnic profile was both helpful and a drawback in the analysis. We cannot use data from this study region to test whether areas with larger ethnic minority populations had more cases or deaths. However, ethnicity not being an unobserved or poorly measured confounder makes it simpler to interpret the other candidate risk factors.

## CONCLUSIONS

The number of cases in LSOAs were clearly dependent on the age demographic of the populations and their lack of ruralness. Older profile and less rural areas had more cases and more deaths. Allowing for the local age structure and rurality, socio-demographic status of the LSOA was linked with deaths from COVID-19, but not full incidence of Pillar 1 cases in the wider study region. The results indicate that deprivation is an important predictor of poor outcomes subsequent to infection.

## Data Availability

The data are not available. They are anonymised but still records of individuals who did not give consent for wide distribution. Our IRB did not include wide distribution.

## Author Contributions

JB conceived of the study. JB and TW obtained datasets for parameters recorded at LSOA level. SR undertook analysis with input from JB. TW generated the maps. JB wrote the first draft and assembled revisions with comments from all authors. All authors have read and approve of the final version of the manuscript.

## Acknowledgements

Sam Weston and Steven Powles (NHS Norfolk and Waveney CCG) collated, cleaned and pseudo-anonymised the data. In turn, their efforts and ours were supported by the many NHS trusts that operate in Norfolk and Suffolk. Pete Best at the Norfolk & Waveney Health & Care Partnership was instrumental in helping us to obtain the dataset. Jon Fox of NWCCG facilitated data access and clarified how many NWCCG patients attend which acute services where. Thanks to Patrick Spragg at Arden & GEM CSU for explaining the types of services offered by participating NHS trusts.

## APPENDICES

## Appendix 1.

These tables show the LSOA percentile values for numeric variables (ratios of local values to hazardous thresholds). Norfolk & Waveney area compared to all England, or the %s within the rural/urban categories. N&W LSOAs are fairly representative of national population exposures for SO_2_, Benzenes and particulate levels. N&W has better total air quality and lower NO_2_. N&W LSOAs rate as slightly more deprived than the national profile; N&W has slightly longer driving times to reach employment centres. N&W is much more rural than most of England, which results in lower population density and less air pollution.

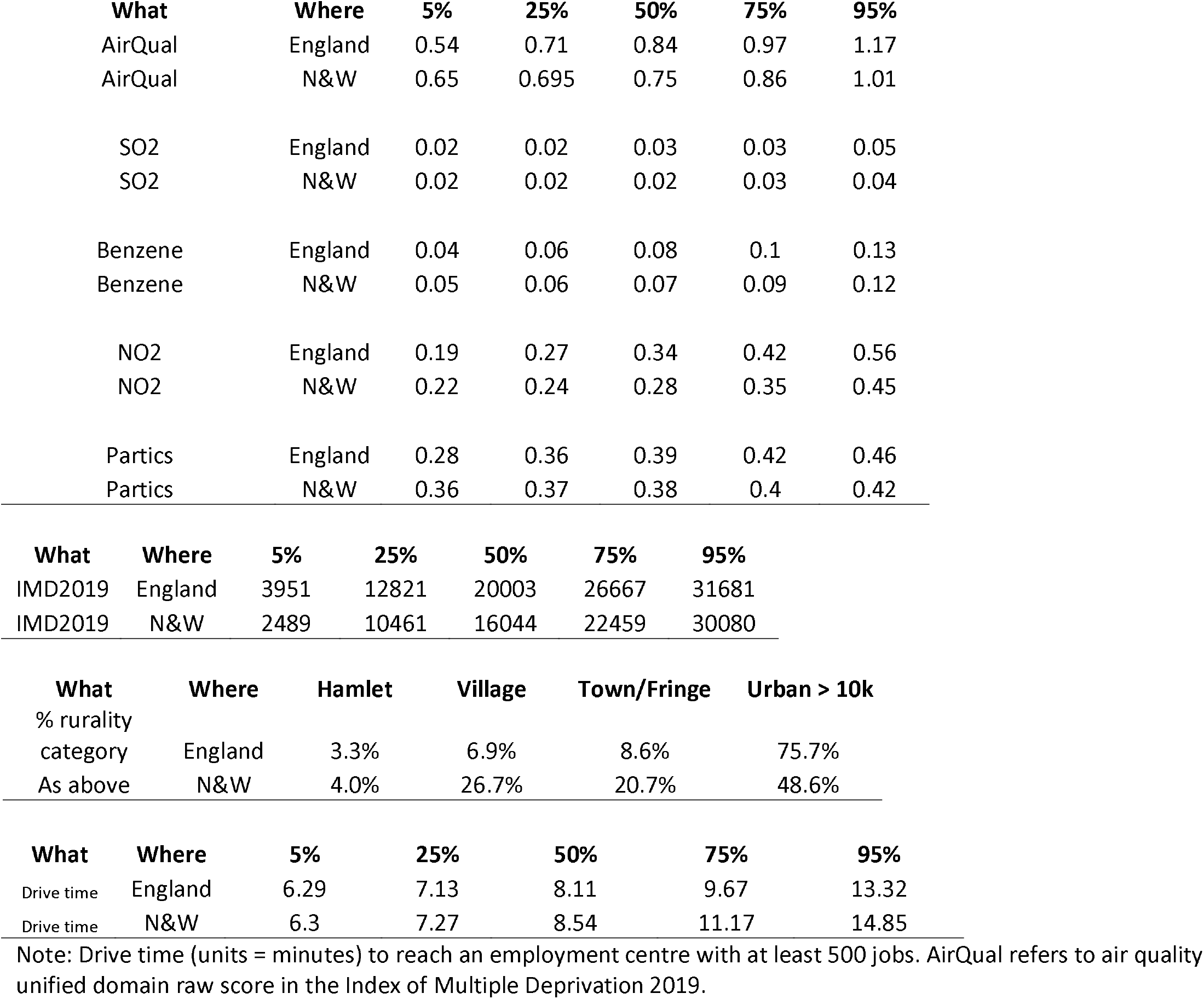

## Appendix 2.

The patients who died were 58% male, 42% female. 1.7% did not have underlying conditions. Deaths may have occurred > 28 days after first +swab test. Deaths occurred in the period 24 March – 30 June 2020. Data come from Norfolk and Norwich University Hospital (NNUH) only. The NNUH is the single largest facility providing acute health care services to the N&W population (see supporting data in Appendix Table 3). 86% of deaths were among persons age 70+. Source: Data published by NNUH at http://www.nnuh.nhs.uk/news/2020/05/daily-announcement-covid-19-deaths/.

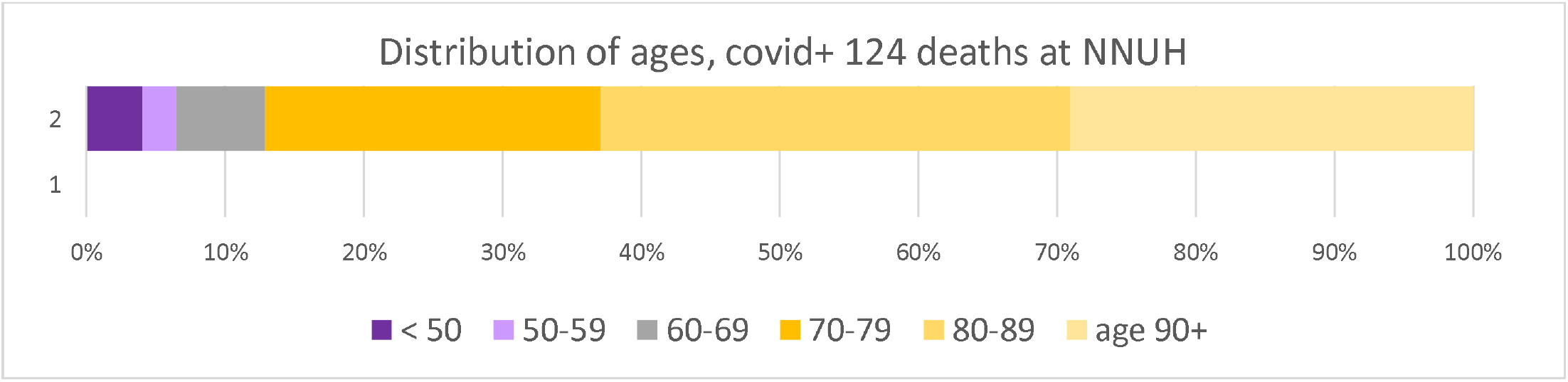

## Appendix 3.

Emergency admissions of Norfolk & Waveney resident patients to acute care providers, aggregated by each of the five constituent clinical commissioning group areas for financial year April 2019-March 2020. Source: Jon Fox of NWCCG, data held as of 22.5.2020.

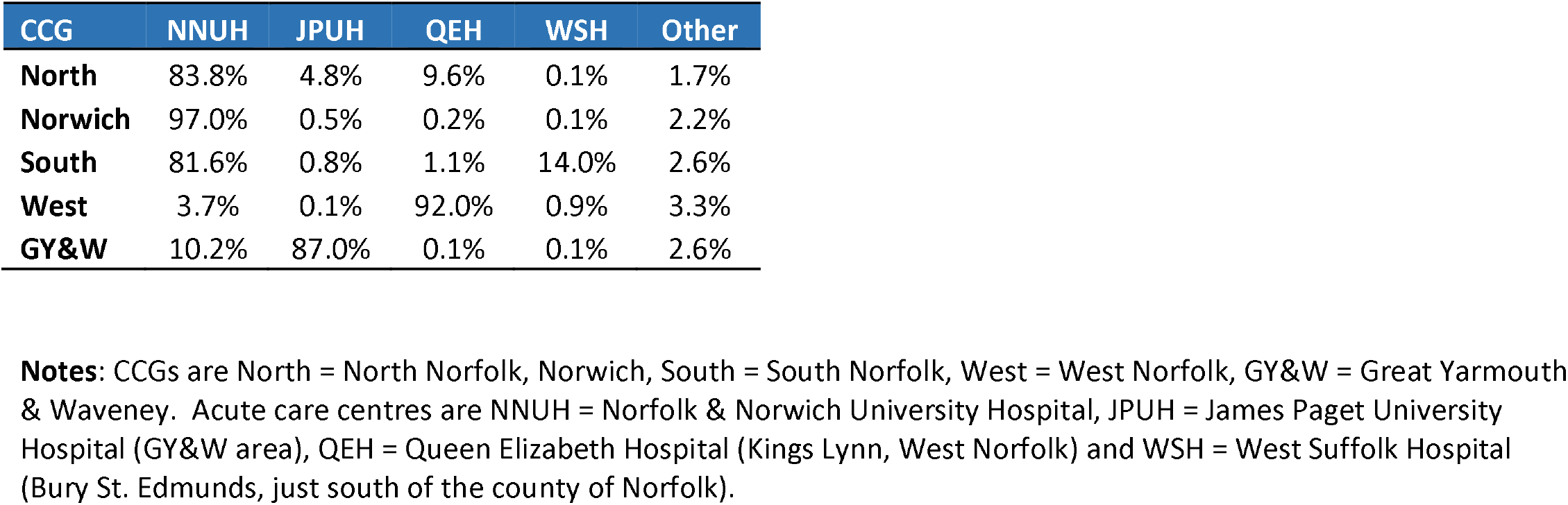

| CCG | NNUH | JPUH | QEH | WSH | Other |
| --- | --- | --- | --- | --- | --- |
| **North** | 83.8% | 4.8% | 9.6% | 0.1% | 1.7% |
| **Norwich** | 97.0% | 0.5% | 0.2% | 0.1% | 2.2% |
| **South** | 81.6% | 0.8% | 1.1% | 14.0% | 2.6% |
| **West** | 3.7% | 0.1% | 92.0% | 0.9% | 3.3% |
| **GY&W** | 10.2% | 87.0% | 0.1% | 0.1% | 2.6% |

**Notes:** CCGs are North = North Norfolk, Norwich, South = South Norfolk, West = West Norfolk, GY&W = Great Yarmouth & Waveney. Acute care centres are NNUH = Norfolk & Norwich University Hospital, JPUH = James Paget University Hospital (GY&W area), QEH = Queen Elizabeth Hospital (Kings Lynn, West Norfolk) and WSH = West Suffolk Hospital (Bury St. Edmunds, just south of the county of Norfolk).

## Appendix 4.

Selection procedure for eligible records.

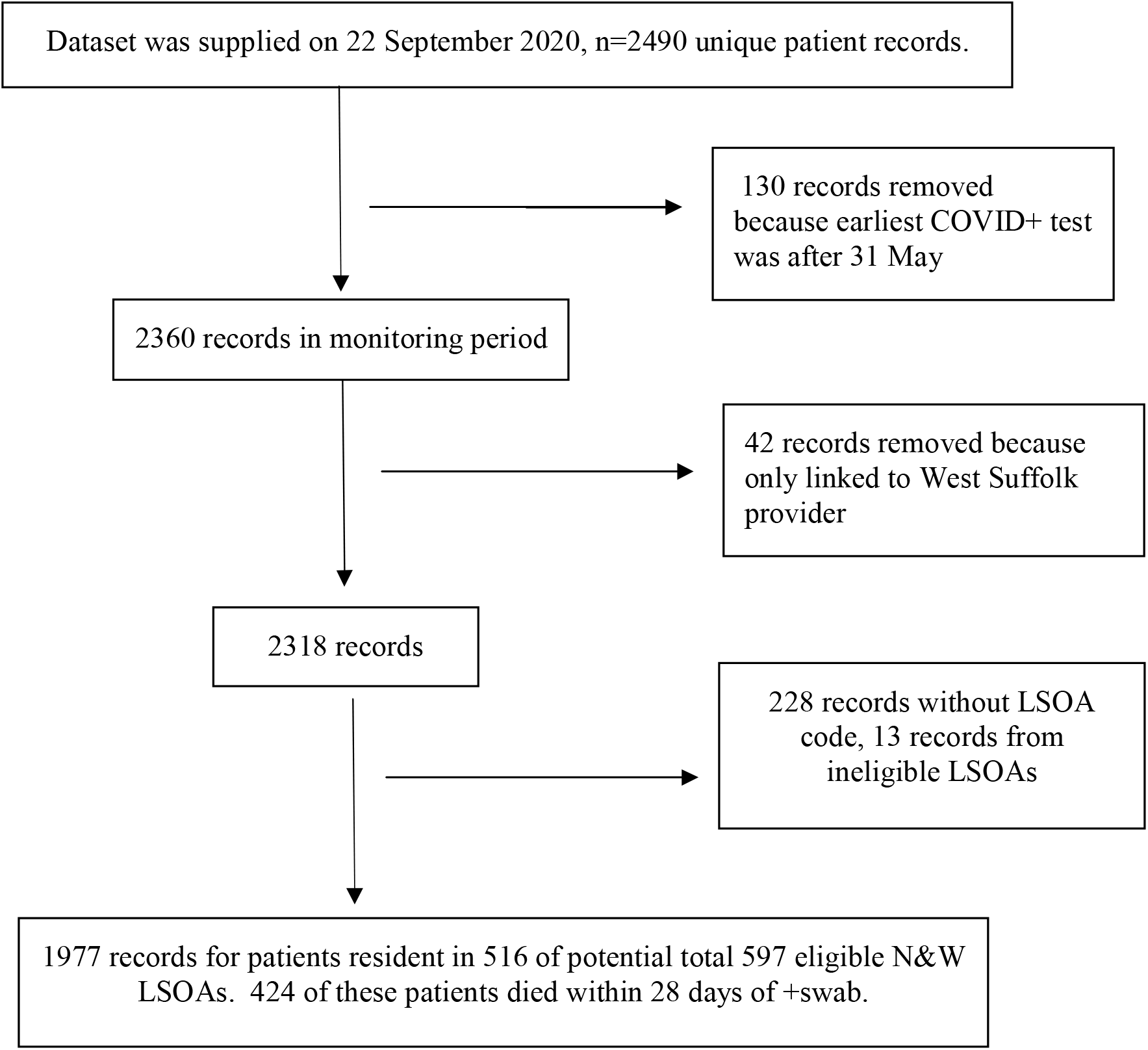

## Appendix 5.

Comparison of demographics of persons in Pillar 1 and Pillar 2 testing frameworks, through 6 August 2020. We include these data to address whether Pillar 2 patients might be very different from Pillar 1. Pillar 2 data were only for residents of the county of Norfolk (we do not have Pillar 2 data for the district of Waveney). The data show some differences in demographic profile of cases found under Pillar 1 vs. Pillar 2 test frameworks.

By 6 August, Fewer persons had tested positive under the Pillar 2 framework than under Pillar 1 sampling frame. The percentage of female persons who tested positive under either Pillar 1 or Pillar 2 were fairly similar. However, males were more likely to be Pillar 1 patients at age 50+. Persons age 50-59 or age 80+ were more likely to be tested under Pillar 1 than Pillar 2. Persons under 20 yrs old were especially likely to be tested under Pillar 2.

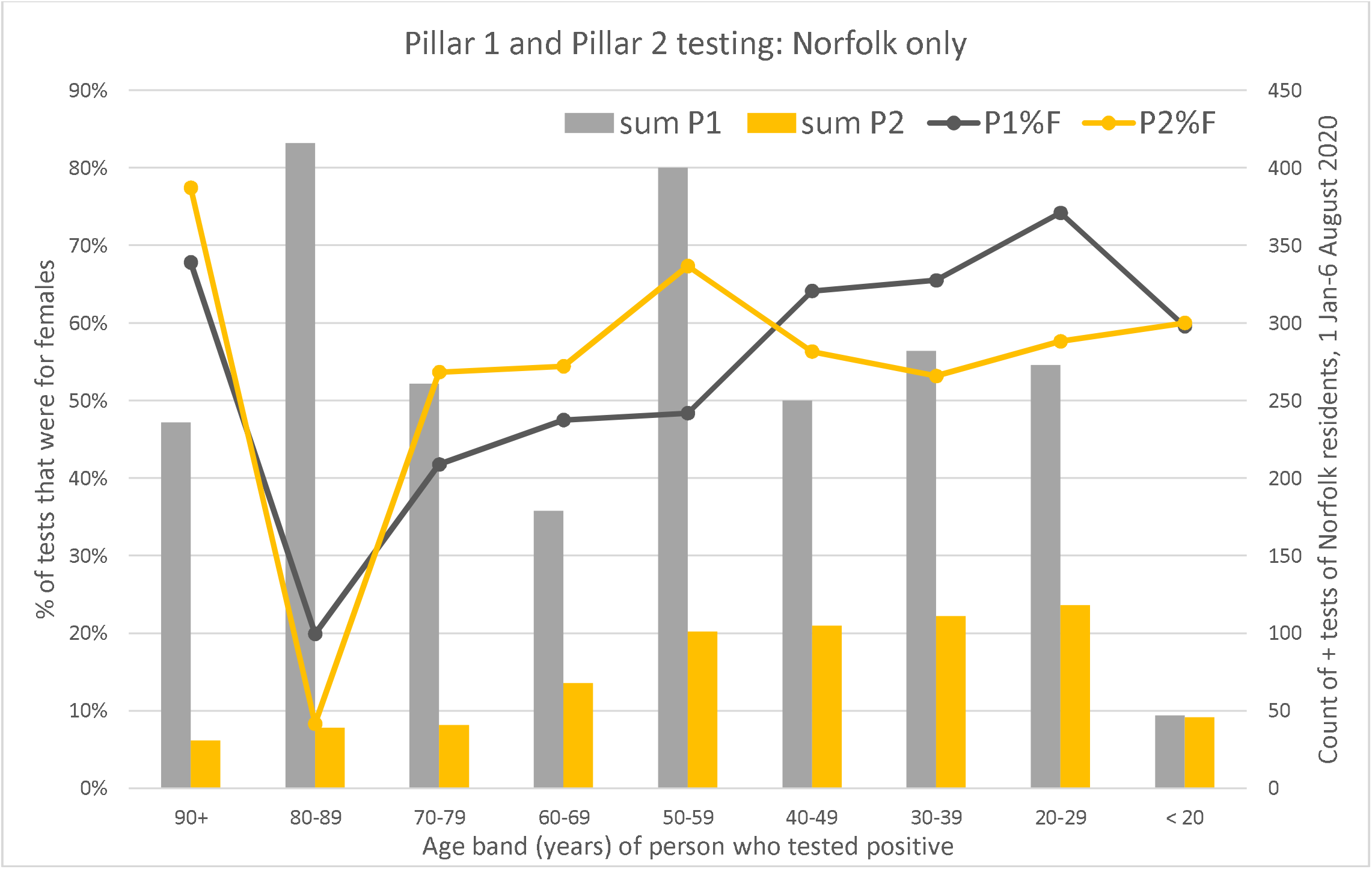

## Appendix 6.

Variance inflation factors for models fit as linear regressions, to test for problematic multi-collinearity that might unduly bias 95% confidence intervals on coefficient estimates.

| Variable | VIF | 1/VIF |
| --- | --- | --- |
| % population age 65+ years | 1.14 | 0.876 |
| Urban rank (higher is more urban) | 1.11 | 0.897 |
| Care home bed capacity | 1.03 | 0.972 |
| Mean VIF | 1.09 |  |

Deaths
| Variable | VIF | 1/VIF |
| --- | --- | --- |
| % population age 65+ years | 1.09 | 0.917 |
| Deprivation rank (higher is less deprived) | 1.06 | 0.939 |
| Care home bed capacity | 1.02 | 0.976 |
| Mean VIF | 1.06 |  |

